# Self-other mentalizing and attachment insecurity in the dimensional model of personality disorders: From research to clinical practice

**DOI:** 10.1101/2025.01.02.25319931

**Authors:** Monika Olga Jańczak, Dominika Górska, Paweł Jurek, Svenja Taubner

**Affiliations:** Faculty of Psychology and Cognitive Science, Adam Mickiewicz University, Poznań, Poland; Institute of Psychology, University of Gdańsk, Gdańsk, Poland; Institute for Psychosocial Prevention, University Hospital Heidelberg, Germany

## Abstract

**Background:** Research on different aspects of mentalizing is essential for understanding the mechanisms underlying personality disorders (PD) and informing psychotherapy approaches, where mentalizing functions as a key mechanism of change. This study aimed to explore whether self- and other-mentalizing, in interaction with attachment insecurity, differentially explain Criteria A (level of personality functioning) and B (maladaptive traits) of the Alternative Model for Personality Disorders in the DSM-5.

**Method:** Our sample consisted of 109 participants (54% female, 41% male, 5% nonbinary). We used The Structured Clinical Interview for DSM-5 Personality Disorders, A Movie for the Assessment of Social Cognition, the Reflective Functioning Questionnaire, the Experiences in Close Relationships – Revised, The Self and Interpersonal Functioning Scale, and The Personality Inventory for DSM-5.

**Results:** Regression analyses show that self-mentalizing deficits uniquely predict both Criterion A domains (self and interpersonal functioning) and all five maladaptive traits, while other-mentalizing is relevant only to interpersonal functioning. Explained variance (adjusted R²) ranges from 55% to 18%. Additionally, mentalization moderates the relationship between insecure attachment and personality pathology (Interpersonal functioning, Negative Affectivity and Detachment), mainly with greater mentalization deficits linked to more severe personality dysfunction under heightened attachment insecurity.

**Conclusion:** Our findings authorize mentalizing as a crucial factor in PD, supporting the potential value of mentalization-focused interventions in addressing both the severity and the “flavor” of PD. Notably, our findings suggest a hierarchy within self- and other-mentalizing, indicating that self-mentalizing plays a more foundational role in PD. Through a comprehensive, multi-method assessment of mentalization, this study offers a refined understanding of its role in psychopathology, providing valuable insights that could guide the development of more targeted therapeutic interventions.

## Introduction

Mentalization is a key factor associated with personality pathology and has been extensively studied in the context of Borderline Personality Disorder (BPD, 1–3) with mentalization-based models of personality disorders (PD) now extending to other diagnoses, such as antisocial and narcissistic PD (4). Recent systematic reviews suggest that mentalizing is a general mechanism of change in psychotherapy, not limited to PD treatment (5). Mentalization is addressed explicitly or implicitly in psychotherapies dedicated to wide range of PD, mainly Mentalization-Based Treatment (MBT; 1), but also Transference-Focused Psychotherapy (TFP) (6,7). Psychotherapy studies often grapple with the complexity of change processes, making conclusion on mediators and moderators of change difficult to pin down (8–10). Recent reviews on mechanisms of change have highlighted the nonspecific nature of many change processes, revealing that they are neither treatment-nor disorder-specific (10,11). Nevertheless, among the numerous proposed mechanisms, mentalizing stands out as one of the most empirically supported, especially in the context of PD (2,12). Meanwhile, a growing body of studies shows that the dimensional, not categorical, diagnosis better explains the pathomechanism of PD, as revealed in a completely new definition of PD in ICD-11 (13) and earlier in an Alternative Model of PD (AMPD), in Criteria A and B, Section III of DSM-5 (14). Recently, level of mentalizing proved to be related both to Criterion A and B of the DSM-5 AMPD (15–17), although the literature on this topic is still scarce and further studies are needed to establish specific relations between mentalizing and new dimensional models of PD.

Fonagy and his team (1) define mentalization as a partially unconscious imaginary mental process of recognition and understanding of one’s own and others’ behavior and experiences, as a result of intentional mental states. However, mentalization is not unidimensional, and one of the key aspects of mentalizing is the self-other dimension. Self-mentalizing refers to the ability to recognize, differentiate, and comprehend causal relationships among one’s own emotions, desires, beliefs, and thoughts. It involves providing a coherent and accurate description of one’s psychological experiences and is essential for self-reflection and emotional regulation (1). Other-mentalizing involves identifying and distinguishing the mental states of others. It requires the ability to infer others’ thoughts, feelings, and intentions independently of one’s own perspective. This capacity is crucial for empathic understanding and effective social interaction, allowing individuals to navigate complex interpersonal relationships (18). Impairments can affect either or both dimensions, leading to distinct manifestations of symptoms depending on which aspect is affected (19–21). Given the multifaceted nature of mentalizing, it is reasonable to advocate for a more nuanced approach that addresses the distinct functions of self- and other-oriented mentalizing in personality disorders (22).

### Mentalizing and Dimensional Diagnosis of Personality Disorders

Although not explicitly stated, mentalizing is one of the foundational concepts underlying Criterion A of AMPD with both mentalization and the level of personality functioning sharing a common developmental framework (17,23). Self-mentalizing is primarily reflected in the Identity Domain (one’s sense of self and emotional regulation through understanding personal mental states) and Self-Direction Domain (awareness of one’s cognitive processes and reflection on personal experiences).). Other-mentalizing is central to the Empathy Domain, which encompasses understanding others’ experiences and motivations, tolerating different perspectives, and recognizing the impact of one’s behavior on others, as well as to the Intimacy Domain, since the capacity for closeness and mutuality requires recognizing the internal states of others. Although the theoretical, developmental, and clinical links between mentalizing and Criterion A domains appear well-founded, research on this topic remains limited and produces mixed results. Studies by Zettl et al. (17), Rishede et al. (16), Maerz et al. (24), and Borroni et al. (25) suggest that all aspects of personality functioning are associated with mentalizing abilities. However, using self-reports or interview-based coding scale, these studies did not specifically differentiate the self-other dimension within mentalizing assessments. Since research suggests that self-report measures tend to emphasize self-oriented mentalizing, it is likely that such studies primarily capture this aspect of mentalization (19). On the contrary, the findings of the Movie for the Assessment of Social Cognition (MASC), which specifically measures the ability to mentalize about others, show different patterns. In the study by Borroni et al. (25), MASC scores were associated only with the interpersonal dimension of personality functioning, while Vizgaitis and Lenzenweger (26) found no significant relationship between MASC scores and any of the Criterion A dimensions. The mixed results concerning the relationship between mentalization and Criterion A may be attributed to the use of diverse assessment methods (19,27) and the specific dimensions of mentalizing measured by different tools (25,26). These findings highlight the need for further research to clarify the relationship between the level of personality functioning and different facets of mentalizing.

While Criterion A of the AMPD is closely linked to mentalization due to their shared theoretical and developmental underpinnings, there is currently no clear theoretical model that directly connects mentalization with the Criterion B traits or explains how mentalization might influence the “flavor” of specific personality disorders (28). Criterion B comprises empirically derived dysfunctional personality domains—Negative Affectivity, Antagonism, Detachment, Disinhibition, and Psychoticism—each defined by associated traits (14). Each of these domains are potentially shaped by the capacity to mentalize, both understanding one’s own and others’ mental states. For instance, in interpersonal contexts, Detachment reflects a withdrawal from relationships, often accompanied by a perception of others as threatening; Antagonism is characterized by hostility toward others, with limited awareness of their needs and feelings (14), whereas Negative Affectivity includes separation anxiety and fear of abandonment and rejection (29). Disinhibition is marked by irresponsibility and a disregard for others, when Psychoticism entails a tendency to attribute thoughts to others or believe one can “read” others’ minds (14). In a self-mentalizing context, Criterion B domains encompass mental states that result from limited self-mentalization, such as anxiety and depression within Negative Affectivity, restricted affect in Detachment, acting outs driven by impulsivity in Disinhibition, and atypical cognitive processes associated with impaired reality testing and weak self-other differentiation in Psychoticism (14).

It seems that maladaptive traits may function as specific vulnerabilities, increasing the likelihood of losing mentalizing capacities in particular trigger situations. This suggests a dynamic process influenced by specific trait combinations that may be observed in patients with PD. However, research on the relationship between Criterion B and mentalization remains limited. Existing studies consistently show that low self-mentalizing is positively associated with certain maladaptive personality traits (30,31). Regarding other-oriented mentalization assessed through the MASC task, significant correlations have been found with traits across all five domains (15). Hanegraaf et al. (32) reported that individuals with high levels of Antagonism and Disinhibition exhibited poorer other-oriented mentalizing compared to those with elevated Detachment and Negative Affectivity. It seems that both self- and other-oriented mentalizing are somehow associated with all maladaptive personality traits. Specifically, other-oriented mentalizing regulates interpersonal strategies, while self-oriented mentalizing “colors” self-perception and self-regulation. However, these findings require replication that gives more deeper and nuanced understanding of how different dimensions of mentalization (i.e., self versus other) interact with personality traits.

### Mentalization and PD in the context of attachment

Research indicate that insecure attachment may contribute to disturbances in mentalizing, leading to intrapersonal and interpersonal difficulties characteristic of PD (5). Although there are only a few studies examining the relationship between insecure attachment and impaired personality functioning within Criterion A of the AMPD (33–35), evidence suggests that attachment anxiety is a stronger predictor of impaired self-functioning, while attachment avoidance is a stronger predictor of impaired interpersonal functioning (36). Studies also show that all Criterion B domains are related to insecure attachment, with the strongest associations found for Negative Affectivity and Detachment, and the weaker connections observed for Disinhibition and Psychoticism (37). In other studies, attachment anxiety emerges as a robust predictor of Negative Affectivity, whereas attachment avoidance is a strong predictor of Detachment (38,39), also in the ICD-11 maladaptive traits model (40). When Relationship Questionnaire is used, fearful attachment shows negative correlations with Negative Affectivity, Detachment and Psychoticism and dismissing attachment is associated with Negative Affectivity and Antagonism (33,34).

Taken together, studies suggest that both mentalizing and attachment influence the level of personality functioning and maladaptive personality traits when examined separately. However, despite the well-established link between mentalization and attachment, they are surprisingly rarely examined together as joint explanatory factors for PD as conceptualized in the DSM-5 AMPD framework. To date, only one study has explored the relationship between attachment and Criterion B in the context of mentalization about others, demonstrating that other-mentalizing moderated the relationship between attachment avoidance and Negative Affectivity, but not for any other maladaptive trait domains (41). The significant role of mentalization in the relationship between attachment and PD has also been supported in research beyond the AMPD framework (e.g., 42). This underscores the need for further exploration of dimensional models of PDs, particularly in examining the interplay of self-other mentalization in the attachment context.

### Aims of the study

This study investigates the relationship between mentalization, attachment, and the AMPD of DSM-5, focusing on both Criterion A (level of personality functioning) and Criterion B (maladaptive personality traits). We propose that addressing different dimensions of mentalization (self vs. other) may advance our understanding of the mechanisms underlying personality disorders within the AMPD framework.

1) For Criterion A, we expect that self-oriented mentalizing will be associated with difficulties in both self- and interpersonal functioning, while other-oriented mentalizing will be linked solely to interpersonal functioning. 2) For Criterion B, we anticipate that both self- and other-oriented mentalizing will be related to maladaptive personality traits, as each trait inherently involves representations of both the self and others specific for this trait. We propose that the associations of self- and other-oriented mentalizing with Criterion B traits are not tied specifically to the „flavour” of the traits but rather to their level of descriptiveness. 3) Additionally, we hypothesize that mentalizing will moderate the relationship between attachment insecurity and personality disorders, functioning as a protective factor against personality pathology in the context of insecure attachment.

To address gaps in previous research, we applied two distinct methods to assess mentalizing: one for self-oriented and another for other-oriented mentalizing, meeting the need for a multimethod approach. To capture individuals with varying levels of personality pathology (none, mild, or severe) and acknowledge that only a subset of individuals with PD seek mental health services, we used two sampling sources: a clinical sample from psychiatric services (inpatients) and community-recruited participants from the local community. This approach enhanced sample heterogeneity in terms of adaptation levels.

## Methods

### Transparency and Openness

We report how we determined our study sample and how we handled missing data, all manipulations, and all measures in the study, and we follow Journal Article Reporting Standards (43). All data, analysis code, and research materials are available online. This study was retrospectively registered, with the analytic plan registered before data analysis, following the guidelines outlined by van den Akker et al. (44); the registration is available at https://osf.io/wzrhn/?view_only=533bff020a6f406d929703e9560ff5b3 [anonymized link]. We detailed how we expanded our analyses compared to our initial statistical plan, as outlined in the Statistical Analyses section.

### Participants and Procedure

A power analysis was conducted using the *pwr* package in R to estimate the minimum required sample size for the multiple regression models, which included five predictors. Assuming a medium effect size *f^2^* = 0.15, a significance level of *p* = 0.05, and a power of 1-beta = 0.80, the analysis indicated that at least 91 observations are required to detect a statistically significant effect. Our sample consisted of 109 participants (54% female, 41% male, 5% nonbinary). Participants was recruited from psychiatric inpatient departments (N = 60), universities, and the local community (N = 49) and were 18 to 47 years of age (M = 25, SD = 6.8). Educational levels varied among participants: 58% had completed secondary education, 35% held university degrees, 4% had vocational training, and 4% had primary education. The community sample comprised individuals who voluntarily participated in a comprehensive onsite psychiatric assessment in response to an invitation to a study on mental health issues and interpersonal functioning. A PD diagnosis was not required for study inclusion, as the study aimed to examine the full range of personality functioning, including cases of subthreshold and absent personality pathology. The exclusion criteria were acute episodes of psychosis, major depression or bipolar disorder, substance dependence, brain injuries, and language difficulties. Participants were compensated for their time with a shopping voucher. The study procedure was carried out in person by clinicians with the experience in therapeutic intervention for patients with severe PD. All participants gave written informed consent for their participation in this research. The study was approved by the Human Research Ethics Committee at [anonymized for review], decision no. 6/07/2021. Data were collected between September 1, 2021, and July 23, 2022.

In the entire sample, 70% had attended psychotherapy at some point of their lives: 57% of the community sample and 82% of the inpatient group, χ^2^(1, *N* = 109) = 7.82, *p =* .005. A total of 54% of the participants met criteria for a PD, according to Section II of DSM-5, assessed with the Structured Clinical Interview for DSM-5 – PDs (77% of the inpatient group and 26% of the community sample). The most prevalent was borderline (30% of the sample), obsessive-compulsive (19%), avoidant (18%), dependent (9%), paranoid (6%), antisocial (6%), narcissistic (4%), histrionic (3%), schizoid (1%) and other (1%). Notably, there was a high level of comorbidity among PD diagnoses, with 23% of the sample having two or more comorbid PD diagnoses (39% of inpatient group and 4% of the community sample).

### Measures

**The Structured Clinical Interview for DSM-5 Personality Disorders** (SCID-5-PD) (45) was employed to assess all 10 personality disorders listed in Section II of the DSM-5. This method assigns categorical diagnoses based on the presence of clinically significant disturbances that meet the specific diagnostic criteria defined in the DSM-5.

**The Movie for the Assessment of Social Cognition** (MASC, 46) is a 15-minute film depicting four people meeting for dinner. Participants are tasked with recognizing the characters’ mental states by answering 46 multiple-choice questions about the characters’ feelings, thoughts, or intentions, primarily focusing on dating and friendship-related issues. Each question offers four response options: (a) a hypermentalizing response, (b) an undermentalizing response, (c) a nonmentalizing response, and (d) a correct mentalizing response. Summary scores for each subscale are calculated based on the frequency of selected responses. Additionally, six control questions are included. Participants have 30 seconds to respond to each question. The MASC demonstrates good reliability, with Cronbach’s α = .73 for the total score.

**The Reflective Functioning Questionnaire** (47) is an 8-item self-report measure assessing mentalization. It includes two subscales: Certainty (RFQ_C) and Uncertainty (RFQ_U) about mental states. Based on prior research (48,49), we used only the RFQ_U subscale in our study, as the validity of the RFQ_C is not well-established, and its specific measurement focus remains unclear. The RFQ_U captures hypomentalization, with higher scores indicating greater difficulties in mentalizing. All items on the RFQ_U refer specifically to mentalizing about oneself. In our study, Cronbach’s α for the RFQ_U was .79.

**The Self and Interpersonal Functioning Scale** (50) is a 24-item self-report questionnaire designed to assess the four core elements of personality pathology outlined in Criterion A of the DSM-5. These elements include two self-related aspects (Identity and Self-direction) and two interpersonal aspects of personality functioning (Empathy and Intimacy), reflecting broader dimensions of Self and Interpersonal functioning. Higher scores indicate more severe impairments in personality functioning. In our study, Cronbach’s alpha reliability coefficients were α = .85 for self-functioning, and α = .82 for interpersonal functioning.

**The Personality Inventory for DSM-5** (51) is a 220-item questionnaire with a 4-point response scale (0 = very false or often false to 3 = very true or often true), which was explicitly designed to measure the proposed DSM-5 Criterion B maladaptive personality traits. The PID-5 has 25 primary scales that load onto five higher order dimensions. In our study, the Cronbach’s alpha values were high for all measured dimensions: Negative Affectivity (α = .96), Detachment (α = .97), Antagonism (α = .95), Disinhibition (α = .91), and Psychoticism (α = .95).

**The Experiences in Close Relationships – Revised** (ECR-R, 52) is a self-report questionnaire designed to assess two dimensions of adult attachment: attachment anxiety (concerns about a partner’s availability and responsiveness) and attachment avoidance (discomfort with closeness and dependency in relationships). Higher scores indicate greater attachment anxiety and/or avoidance, while low scores on both dimensions reflect secure attachment. In our study, the reliability of the ECR-R was very high, with Cronbach’s α = .93 for attachment anxiety and α = .90 for attachment avoidance.

### Statistical analyses

All calculations were conducted using the R programming environment (53), supplemented by relevant statistical packages. To handle missing data, we applied the following procedures: for questionnaires scored using sum totals, missing values were replaced with the scale’s average score; for questionnaires scored using mean values, missing data points were excluded. The percentages of missing data were 0.74% for PID-5, 1.53% for SIFS, 9.29% for RFQ-8, 0.15% for ECR-R, and 1.55% for MASC. The higher rate of missing data for RFQ-8 resulted from the first study group not receiving this questionnaire. No missing data were reported for SCID-5-PD.

After inspecting the data, we assessed the internal consistencies of the study measures using Cronbach’s alpha (α). Coefficients of at least .65 were deemed acceptable, while values of .70 or higher indicated good internal consistency. All measures met satisfactory reliability standards (αs ≥ .67; see Measures for details). Descriptive statistics were then calculated to summarize the data. To assess the univariate normality of variable scores, we applied cut-off criteria for skewness and kurtosis between −2 and +2, which indicated minimal departure from normality. Consequently, the distributions of all continuous variables in our study were fairly symmetrical, and parametric statistics were used.

Next, we computed correlations to explore relationships between the study variables. Pearson’s r was used for correlations between continuous variables. To test the hypotheses that various facets of mentalizing are associated with dimensions of personality pathology, a series of multiple regression analyses were performed. Seven regression models were used to explore the relationships between self- and other-focused mentalizing and different aspects of personality disorders. This strategy was selected to quantify the unique contribution of each mentalizing facet to the dimensions of personality pathology. The predictors included five distinct facets of mentalizing: one scale for self-mentalizing and four for other-mentalizing (hypermentalizing, non-mentalizing, undermentalizing, and correct mentalizing). The dependent variables comprised self- and interpersonal functioning (Criterion A) and the five maladaptive personality traits: Negative Affectivity, Antagonism, Detachment, Disinhibition, and Psychoticism (Criterion B). Variance Inflation Factor (VIF) scores were calculated for the predictor set to assess multicollinearity, with all predictors demonstrating acceptable VIF values. Standardized regression coefficients (β) were interpreted to evaluate the strength and direction of the associations, adjusted R² values were used to assess the explanatory power of each model, and p-values were examined to determine the statistical significance of the observed relationships.

Another series of regression models was conducted to examine the moderating effects of mentalizing on the relationships between attachment insecurity and personality pathology (seven dependent variables for both Criterion A and Criterion B). For each of the dependent variables, five regression models were tested. The moderating roles of five distinct mentalizing facets were assessed: self-mentalizing and four aspects of other-mentalizing (correct mentalizing, hypermentalizing, undermentalizing, and non-mentalizing). These facets were examined in relation to anxious and avoidant attachment. In total, 35 models were tested. To enhance interpretability, the independent variables involved in interaction terms were mean-centered prior to analysis.

In our preregistration, we initially planned to conduct a mediation analysis using Structural Equation Modeling (SEM) rather than performing a series of regression and moderation analyses. These changes were made because the revised approach aligned better with our data.

## Results

### Regression Analysis for Criterion A and Criterion B of AMPD

Self-mentalizing deficits were strongly associated with both the Self and Interpersonal functioning dimensions of Criterion A and showed moderate to strong associations with all Criterion B traits (see Table 1). Furthermore, strong positive correlations were observed between self-mentalizing deficits and attachment anxiety. Other-hypermentalizing was weakly to moderately associated with Antagonism, Psychoticism, and, to a lesser extent, with Negative Affectivity and Disinhibition. Correct other-mentalizing scores showed weak negative correlations with the Interpersonal functioning of Criterion A and negligible associations with attachment anxiety and avoidance. No significant correlations emerged between correct other-mentalization and Criterion B traits.

**Table 1.** Correlations between study variables.

|  | Self-functioning difficulties | Interpersonal functioning difficulties | Negative Affectivity | Detachment | Antagonism | Disinhibition | Psychoticism | Self-mentalizing deficits | Correct mentalizing | Hiper-mentalizing | Under-mentalizing | Nonmentalizing | Anxious attachment |
| --- | --- | --- | --- | --- | --- | --- | --- | --- | --- | --- | --- | --- | --- |
| Interpersonal functioning difficulties | 0.62*** | — |  |  |  |  |  |  |  |  |  |  |  |
| Negative Affectivity | 0.781*** | 0.565*** | — |  |  |  |  |  |  |  |  |  |  |
| Detachment | 0.705*** | 0.617*** | 0.707*** | — |  |  |  |  |  |  |  |  |  |
| Antagonism | 0.398*** | 0.569*** | 0.532*** | 0.237* | — |  |  |  |  |  |  |  |  |
| Disinhibition | 0.534*** | 0.269** | 0.297** | 0.162 | 0.352*** | — |  |  |  |  |  |  |  |
| Psychoticism | 0.645*** | 0.431*** | 0.621*** | 0.467*** | 0.534*** | 0.514*** | — |  |  |  |  |  |  |
| Self-mentalizing deficits | 0.749*** | 0.508*** | 0.727*** | 0.443*** | 0.552*** | 0.499*** | 0.635*** | — |  |  |  |  |  |
| Correct mentalizing | -0.147 | -0.353*** | -0.128 | -0.154 | -0.187 | -0.037 | -0.101 | -0.072 | — |  |  |  |  |
| Hipermentalizing | 0.252** | 0.187 | 0.244* | 0.119 | 0.335*** | 0.205* | 0.323*** | 0.310** | -0.498*** | — |  |  |  |
| Undermentalizing | -0.146 | 0.034 | -0.157 | -0.034 | -0.004 | -0.081 | -0.123 | -0.185 | -0.414*** | -0.059 | — |  |  |
| Nonmentalizing | -0.096 | 0.071 | -0.066 | 0.023 | -0.019 | -0.062 | -0.058 | -0.209* | -0.524*** | 0.137 | 0.290** | — |  |
| Anxious attachment | 0.700*** | 0.594*** | 0.755*** | 0.506*** | 0.504*** | 0.386*** | 0.537*** | 0.640*** | -0.190* | 0.237* | -0.043 | -0.043 | — |
| Avoidant attachment | 0.280** | 0.441*** | 0.109 | 0.550*** | 0.107 | 0.039 | 0.164 | 0.114 | -0.221* | 0.077 | 0.137 | 0.097 | 0.169 |

Note: \* $p < .05$ , \*\* $p < .01$ , \*\*\* $p < .001$ ; Mentalization about self: Self-mentalizing deficits; Facets of mentalization about others: Correct mentalizing, Hipermentalizing, Undermentalizing, Nonmentalizing

Table 2 shows the results of multiple regression analyses. Correct other-mentalizing significantly predicted interpersonal functioning difficulties (β = −0.43, p < .01). The strongest predictor across all models was self-mentalizing, which had a large positive effect on self-functioning difficulties (β = 0.75, p < .01), interpersonal functioning difficulties (β = 0.52, p < .01), and Negative Affectivity (β = 0.74, p < .01). The rest of other-mentalizing dimensions, such as hypermentalizing, undermentalizing, and nonmentalizing, showed no significant predictive effects for most outcomes. The models explained between 18% and 55% of the variance in the dependent variables, as reflected by the adjusted R² values. The highest variance explained was for self-functioning difficulties (R² = .55) and Negative Affectivity (R² = .52).

**Table 2.**
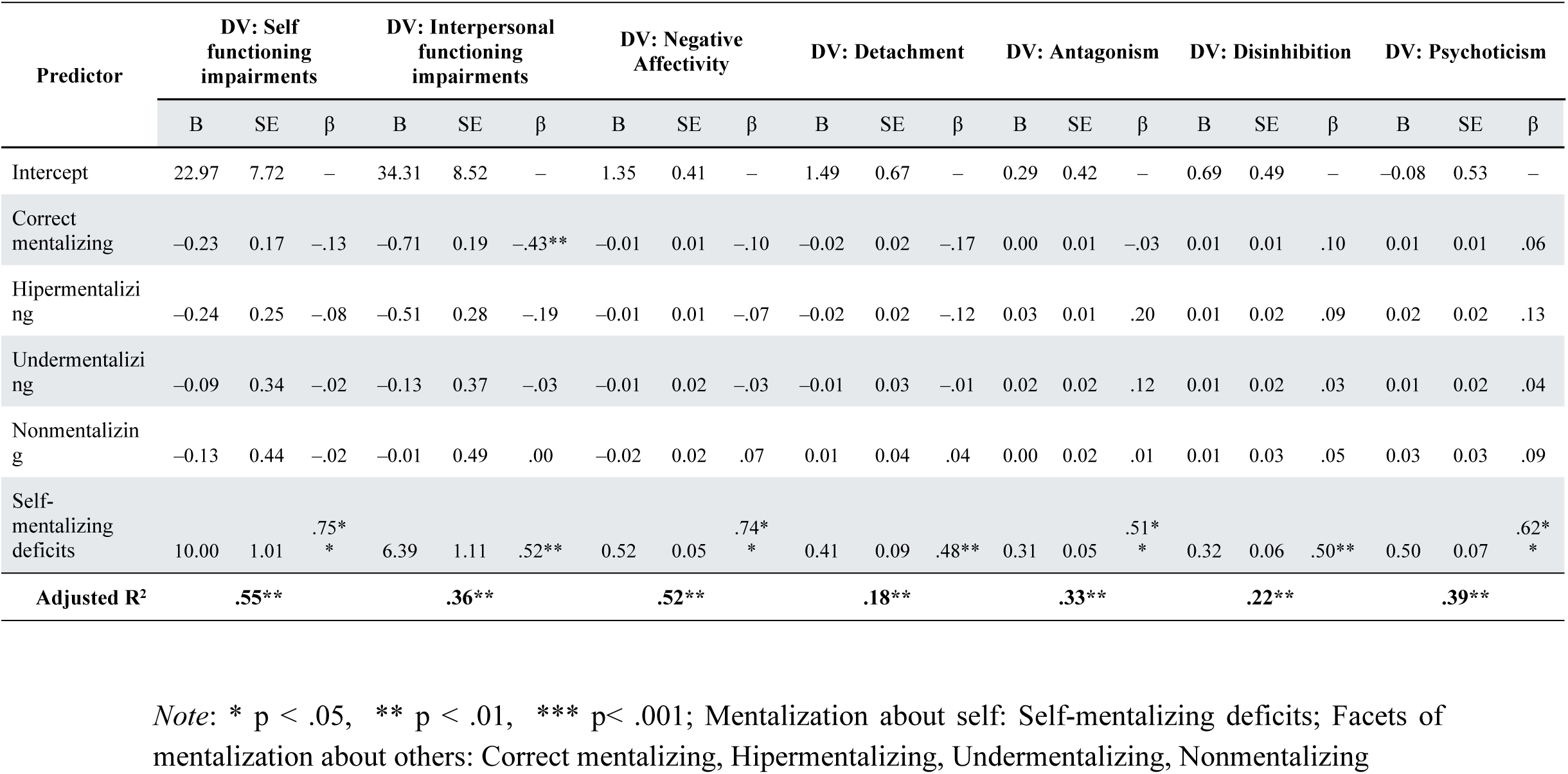
Regression Analysis for Dependent Variables: Level of Personality Functioning and Maladaptive Trait.

### Moderation Analysis for Effects of Self- and Other-Mentalizing on the Relationship Between Attachment and Criterion A and B

The results of the moderation analysis for the prediction of each dependent variable are presented in the Supplementary Materials (Tables S1 to S7, Figs S1-S3), with each table dedicated to a specific dimension of personality pathology. Table 3 summarizes the significant effects identified in the analyses.

**Table 3.** Summary of Significant Interaction Effects in Regression Analyses.

| Dependent Variable |  | Moderator |  |  |  |  |
| --- | --- | --- | --- | --- | --- | --- |
|  |  | Self-mentalizing | Other correct mentalizing | Other hypermentalizing | Other undermentalizing | Other nonmentalizing |
| Self-functioning difficulties | Main effects | ANX, AVO, Self-mentalizing | ANX, AVO | ANX, AVO | ANX, AVO, Undermentalizing | ANX, AVO |
|  | Interaction effects | - | - | - | - | - |
| Interpersonal functioning difficulties | Main effects | ANX, AVO, Self-mentalizing | ANX, AVO | ANX, AVO | ANX, AVO | ANX, AVO |
|  | Interaction effects | - | Correct mentalizing x ANX | Hypermentalizing x ANX | - | - |
| Negative Affectivity | Main effects | ANX, Self-mentalizing | ANX | ANX | ANX | ANX |
|  | Interaction effects | Self-mentalizing x ANX | - | - | - | - |
| Detachment | Main effects | ANX, AVO, Self-mentalizing | ANX, AVO | ANX, AVO | ANX, AVO | ANX, AVO |
|  | Interaction effects | Self-mentalizing x ANX | Correct mentalizing x AVO | - | Undermentalizing x AVO | Nonmentalizing x AVO |
| Antagonism | Main effects | ANX, Self-mentalizing | ANX | ANX, Hypermentalizing | ANX | ANX |
|  | Interaction effects | - | - | - | - | - |
| Disinhibition | Main effects | Self-mentalizing | ANX | ANX | ANX | ANX |
|  | Interaction effects | - | - | - | - | - |
| Psychoticism | Main effects | ANX, Self-mentalizing | ANX | ANX, Hypermentalizing | ANX | ANX |
|  | Interaction effects | - | - | - | - | - |

*Note:* ANX - anxious attachment, AVO - avoidant attachment

In the models predicting Criterion A self-functioning impairments, both anxious attachment and avoidant attachment emerged as significant predictors (see Table S1, Supplementary Materials). However, no interaction effects with mentalizing were observed.

In the models predicting Criterion A interpersonal functioning impairments, both attachment scales generally emerged as significant predictors (see Table S2, Supplementary Materials). Additionally, we observed moderation effect: Mentalizing about others alters the effect of attachment anxiety on interpersonal functioning impairments. Correct other-mentalizing significantly moderated the relationship between anxious attachment and interpersonal functioning (B = −3.33, SE = 0.08, Beta = −0.31, p < 0.01) (see Fig S1a, Supplementary Materials). The interaction effect indicated that among individuals with higher levels of correct other-mentalizing, the relationship between anxious attachment and interpersonal functioning was significantly weaker. Among individuals with high levels of anxious attachment, those with higher levels of correct mentalization exhibit lower interpersonal functioning impairment compared to those with lower levels of correct mentalization. Other-hypermentalizing also significantly moderated the relationship between anxious attachment and interpersonal functioning (B = 0.32, SE = 0.14, Beta = 0.17, p < 0.05) (see Fig S1b, Supplementary Materials). This interaction effect showed that among individuals with lower levels of hypermentalizing about others, the relationship between anxious attachment and interpersonal functioning impairment was significantly weaker. Among individuals with high attachment anxiety, those with higher levels of hypermentalization about others exhibit greater levels of interpersonal functioning impairments compared to those with lower other-hypermentalization.

In the model predicting Negative Affectivity, Detachment, Antagonism, Disinhibition, and Psychoticism, anxious attachment generally emerged as significant predictors (see Table S3-S7, Supplementary Materials). Moreover, we observed moderation effect: Mentalizing about self reduces the effect of attachment anxiety on both Negative Affectivity and Detachment. Self-mentalizing significantly moderated the relationship between anxious attachment and NA (B = −0.08, SE = 0.03, Beta = −0.17, p < 0.05). The interaction effect indicated that among individuals with higher levels of self-mentalizing deficits, the relationship between anxious attachment and Negative Affectivity was significantly weaker. Participants with low attachment anxiety and low levels of self-mentalization reported higher Negative Affectivity compared to individuals with similar attachment anxiety scores but better self-mentalization (Fig S2, Supplementary Materials). In the model predicting Detachment, self-mentalizing significantly moderated the relationship between anxious attachment and Detachment (B = −0.10, SE = 0.05, Beta = −0.18, p < 0.05). The interaction effect suggested that among individuals with higher levels of self-mentalization deficits, the relationship between anxious attachment and Detachment was significantly weaker. For individuals with low attachment anxiety, those with greater deficits in self-mentalization exhibited higher levels of Detachment compared to individuals with better self-mentalization (Fig 1).

**Fig 1.**
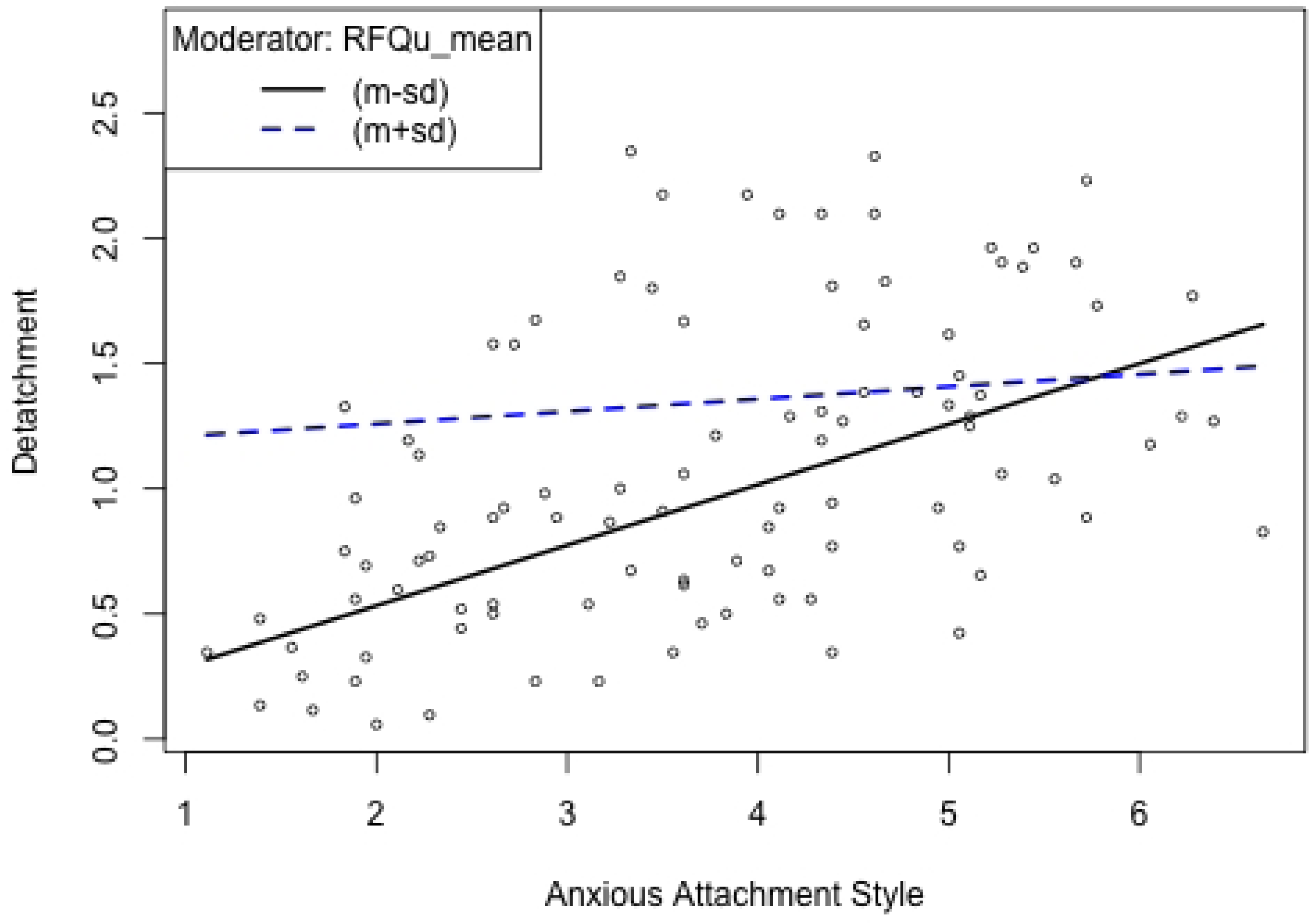
Mentalization about self moderates relationship between Anxious attachment and Detachment. *Note*: RFQu_mean - Self mentalization deficits - Uncertainty

In predicting the five Criterion B traits, a main effect of avoidant attachment was demonstrated only for Detachment. Moreover, we observed a moderation effect for three facets of other mentalizing - they alter the strength of relationship between attachment avoidance and Detachment (S3a—3c Figs, Supplementary Materials). Correct other-mentalizing significantly moderated the relationship between avoidant attachment and Detachment (B = −0.03, SE = 0.01, Beta = −0.21, p < 0.05). This interaction effect revealed that among individuals with higher levels of correct mentalizing, the relationship between avoidant attachment and Detachment was significantly weaker. Other-undermentalizing also emerged as a significant moderator of the relationship between avoidant attachment and Detachment (B = 0.05, SE = 0.02, Beta = 0.16, p < 0.05). The interaction effect showed that among individuals with higher levels of other-undermentalizing, the relationship between avoidant attachment and Detachment was significantly stronger. Similarly, other nonmentalizing significantly moderated the relationship between avoidant attachment and Detachment (B = 0.08, SE = 0.03, Beta = 0.19, p < 0.01). This interaction effect indicated that among individuals with higher levels of other no-mentalizing, the relationship between avoidant attachment and Detachment was significantly stronger. Among individuals with high attachment avoidance, those with greater deficits in other-mentalization exhibit higher levels of Detachment compared to those with better other-mentalization.

## Discussion

Previous research has established that mentalization plays a crucial role in personality disorders (2,5). Studies on mentalization have advanced the understanding of this key mechanism of change in the treatment of PD patients, demonstrating its role as a predictor, moderator, mediator, and outcome variable in psychotherapy research (5). Investigating mentalization aligns with the growing proposition to explore meta-models of change in psychotherapy, offering a framework for studying transtheoretical processes at the mechanism level rather than focusing solely on treatment models (8,10). Our findings further reveal nuanced relationships between different facets of mentalization, attachment insecurity, and various dimensions of personality pathology.

It seems that while self-related mentalization has a prominent, broad and nonspecific predictive role for PD, other-related mentalization is notably more specific, being limited to selected aspects of PD primarily tied to impaired social relationships. Correlation analyses indicate predominantly large effect sizes for self-mentalizing, showing strong associations with both the self and interpersonal domains of personality functioning (Criterion A) and all five maladaptive traits (Criterion B). Other-mentalizing demonstrates small to medium effect sizes, with correct mentalization linked to Criterion A’s interpersonal domain and hypermentalization associated with Criterion A’s self domain and all maladaptive traits except Detachment. Regression analyses further reveal that the significance of other-mentalizing diminishes when self- and other-mentalizing are examined together: deficits in self-mentalizing consistently act as a unique predictor for both aspects of personality functioning and all maladaptive traits, while correct other-mentalizing remains related only to interpersonal functioning in Criterion A. Therefore, we replicated previous findings demonstrating the relationship between mentalization and Criterion A (16,17,24,25), using an economical self-report and performance-based measure of mentalizing while offering a more detailed perspective on the distinction between self- and other-oriented mentalizing. Nevertheless, the percentage of explained variance for self-mentalizing varies significantly. It exceeds 50% for self-functioning and Negative Affectivity, but drops to 18% for Detachment, and 22% for Disinhibition. Theoretically, all aspects of personality pathology involve some degree of representations of self, with Criterion A reflecting overall maturity levels and Criterion B emphasizing distinct “flavors” of personality characteristics (28). Both Criterion A self-functioning and Negative Affectivity primarily involve the awareness and understanding of internal mental states, indicating a strong association with self-mentalizing. In the context of self-functioning, this includes aspects such as self-identity and personal boundaries, while in Negative Affectivity, it pertains to the experience and regulation of emotions (14). The lower explanatory power of self-mentalizing for Detachment and Disinhibition highlights the greater relevance of additional factors influencing these traits. Detachment is characterized by withdrawal from relationships due to perceived interpersonal threats, while Disinhibition reflects maladaptive social behavior driven by temperamental impulsivity and difficulties in regulating behavioral responses (14,54). In summary, while self-related mentalization demonstrates predictive utility across all facets of personality pathology, albeit with varying degrees of explained variance, other-related mentalization exhibits a more specific association with interpersonal dimension, providing nuanced insights that can inform the prioritization and tailoring of treatment objectives we will further discuss.

Moderation analyses revealed a significant main effect of attachment anxiety and avoidance was observed for Criterion A, while for Criterion B, anxiety emerged as significant across all traits, with avoidance showing a main effect only for Detachment. These findings underscore the critical yet distinct role of attachment dimensions in the manifestation of personality disorders, aligning with previous research discussed earlier (33,35,38,39). We also observed moderation effects of self- and other-mentalization on the relationship between insecure attachment and both Criterion A and Criterion B. Mentalization moderates the link between insecure attachment and maladaptive traits such that individuals with greater mentalization deficits show higher levels of personality pathology in the context of elevated insecure attachment. For Criterion A two moderation effects was observed: Individuals with lower levels of mentalization about others (lower correct mentalization and higher hypermentalization) exhibit greater interpersonal difficulties when reporting high levels of anxious attachment. No moderation effect were observed for self-mentalization or for self-functioning difficulties in Criterion A. For Criterion B, other-mentalizing moderates the relationship between avoidant attachment and Detachment, while self-mentalizing moderates the relationship between anxious attachment and both Negative Affectivity and Detachment. These findings diverge somewhat from the results of Ball Cooper et al. (41), who identified a moderation effect of the overall MASC scale between avoidant attachment and Negative Affectivity, but not Detachment. This discrepancy may be attributed to sample differences, as clinical and community samples tend to differ substantially in the severity of maladaptive traits (55). While our study focused on a clinical population, Ball Cooper’s research involved college psychology students, suggesting potentially distinct underlying mechanisms. Nonetheless, the current findings emphasize the pivotal role of both Negative Affectivity and Detachment in the mentalizing-attachment dynamic, so we will discuss that below in more detail.

Both Detachment and Negative Affectivity reflect facets of emotional dysregulation, yet they represent opposing strategies for regulating interpersonal distance. Negative Affectivity is associated with separation anxiety and fear of abandonment, while Detachment involves emotional withdrawal and avoidance of close relationships (14,29). In our study, attachment anxiety was directly associated with Negative Affectivity, with this relationship mitigated by fewer deficits in self-mentalizing. In contrast, Detachment was related to both anxious and avoidant attachment, with mentalization serving as a moderating factor. Specifically, lower mentalization deficits acted as a protective factor, buffering against the severity of Detachment. This buffering effect, however, followed distinct patterns: for anxious attachment, self-focused mentalization moderated the link with Detachment, while for avoidant attachment, other-focused mentalization played this role. These findings highlight the differentiated roles of mentalization polarities in mitigating attachment-related vulnerabilities in personality functioning. They can be discussed in light of Blatt’s (56) two-polarities model of personality, which posits two dimensions: relatedness (anaclitic vulnerability) and self-definition (introjective vulnerability). Relatedness corresponds to anxious attachment and is characterized by excessive preoccupation with relationships and fear of rejection, whereas self-definition aligns with avoidant attachment and reflects discomfort with closeness and dependence on others (57). Our study indicates that mitigating Detachment and Negative Affectivity in individuals with anxious attachment may be facilitated by strengthening self-mentalizing. This probably reduces sensitivity to self-perceived rejection and helps them better regulate relational anxiety. For those with avoidant attachment, on the other hand, better other-mentalizing weakens sensitivity to negative intentions attributed to others, reducing withdrawal and avoidance in close relationships. These nuanced pathways highlight the importance of targeted mentalization strategies to address detachment across different attachment strategies.

### Clinical implications

This study has several clinical implications that could offer a promising foundation for future exploration in psychotherapy research. Our findings further confirm that mentalization is a crucial correlate of personality disorders, underscoring its importance among various psychopathological factors associated with PD. They also support the notion that mentalization-focused interventions may be particularly effective in inducing change in both the severity and ‘flavor’ of personality disorders, extending well beyond mere improvements in social competence. Moreover, our results suggest a hierarchy within self- and other-mentalization polarities, indicating that self-mentalization may play a more foundational role in PD than mentalization directed toward others. This aligns with findings suggesting that successful psychotherapy often begins with strengthening self-mentalization (58). Developing self-mentalization fosters a more differentiated and coherent self-perception, which subsequently enables the capacity to mentalize others by exploring their mental states and motivations. Once self-mentalization is established, individuals can flexibly shift between self- and other-mentalization, facilitating a more integrated and adaptive understanding of interpersonal dynamics. This process reflects transitions from I-mode to me-mode to we-mode observed in MBT (59), and supporting the maintenance of a coherent self-image, reducing splitting, and enabling the exploration of transference roles within the framework of TFP (60). These mechanisms underscore the interplay between intrapersonal and interpersonal mentalizing capacities as foundational elements in the therapeutic resolution of pathological personality structures. Consequently, targeting self-mentalizing deficits should be prioritized in psychotherapeutic interventions for personality disorders, as this appears to be the most important predictor of PD. Future studies should investigate how changes in self- and other-mentalizing unfold during treatment, as well as the specific mechanisms by which improvements in self-mentalization contribute to broader therapeutic outcomes. This psychotherapy processes studies may provide critical insights into optimizing mentalization-based approaches and tailoring interventions to address individual patient needs.

While self-focused mentalization plays a broad role, influencing personality functioning and all traits, other-focused mentalization also holds significant importance but operates in a more specific capacity, targeting particular traits primarily linked to social relationships. Other research show that difficulties in social connectedness are a significant challenge in PD, especially in BPD (61), and that a sense of belonging is a key indicator of recovery outcomes in individuals with PD (62). Our findings suggest that enhancing mentalization about others can specifically address interpersonal aspects of personality, including difficulties in interpersonal functioning (Criterion A) and detachment (regulating interpersonal distance, Criterion B). Developing mentalization skills focused on others may be particularly relevant for individuals who, despite progress in self-related recovery, continue to experience challenges in re-engaging socially.

Additionally, our findings may contribute to understanding the relationships between facets of mentalization, different attachment dimensions, and personality traits in the context of therapeutic relationship. When anxious attachment is activated in patients with high Negative Affectivity, focusing on their internal mental states may be beneficial. Conversely, when avoidant attachment associated with detachment and relational withdrawal is triggered, emphasizing mentalization about others could help reduce withdrawal—potentially lowering the risk of dropout and strengthening the therapeutic alliance. In the light of Blatt’s two polarities theory (57), patients with a predominance of relatedness may benefit from developing the ability to understand and reflect on their own emotions and thoughts, while those with a predominance of self-definition could benefit from enhancing their capacity to infer and interpret the mental states of others for better social understanding.Thus, our study offers a nuanced perspective, providing valuable insights into which aspects of mentalization should be prioritized in therapeutic interventions for specific patient groups.

## Limitations

This study had several limitations. First, the cross-sectional design does not allow for conclusions about the predictive value of the variables over time; a longitudinal approach is needed to establish causal relationships and track changes in mentalizing and personality pathology over time. However, our correlational findings provide a solid starting point for designing more conclusive psychotherapy studies that test moderators and mediators of change in therapies for PD, showing that it is reasonable to distinguish between self- and other-focused mentalization to better understand therapeutic mechanisms and optimize intervention strategies. Secondly, factors beyond personality pathology, such as comorbid symptomatic disorders, were not assessed, which limits the generalizability of the results. Generalizability is further constrained by the heterogeneous sample, which includes both inpatients and community participants. Additionally, differences in assessment methods may have influenced the findings, potentially reflecting method effects rather than true differences in mentalizing dimensions. Future research should address this by comparing self- and other-oriented mentalizing using the same types of methods to improve validity and reduce potential biases. A small sample size is another limitation, suggesting the need for replication with a larger sample. Additionally, to explore specificity for PD, comparisons between individuals with PD, those with other psychiatric conditions, and healthy controls would be valuable. Moreover, while our study employs a nomothetic model, a person-centered approach could be particularly relevant as it may reveal distinct profiles of mentalizing abilities within individuals that differentially relate to personality functioning and maladaptive personality traits. Future studies should address this by clustering mentalizing profiles to create an individualized understanding of these dynamics (20,21). In summary, our findings should be interpreted cautiously given these limitations, and the conclusions are primarily applicable to the specific sample studied.

## Conclusions

This study aims to clarify the complex relationships between mentalizing, attachment, and personality pathology, thereby advancing our understanding of the mechanisms underlying personality disorders within the AMPD framework. Our findings provide insight into why mentalizing is critical in psychopathology and psychotherapy of PD. We proposed that different dimensions of mentalizing (self-oriented vs. other-oriented) would relate to personality pathology in unique ways, especially when considering insecure attachment. Through a comprehensive, multi-method assessment of mentalization, this study provides a more refined understanding of its role in psychopathology, offering insights that may inform mechanisms of change and guide the development of more targeted therapeutic interventions.

## Data Availability

All files supporting this study are available at DOI:(osf link to be added)

https://osf.io/5tvmy/

## Acknowledgments

We sincerely thank all participants for their time and engagement in this study.

## Supporting information

**S1a Fig. Criterion A. Correct mentalization about others moderates relationship between Anxious attachment and Interpersonal functioning.** MASC_correct - correct mentalization about others.

**S1b Fig. Criterion A. Hypermentalizing about others moderates relationship between Anxious attachment and Interpersonal functioning.** MASC_Hiper - Hypermentalizing about others.

**S2 Fig. Criterion B. Mentalization about self moderates relationship between Anxious attachment and Negative Affectivity.** RFQu_mean - Mentalization about self - Uncertainty.

**S3a Fig. Criterion B. Mentalization about others (correct mentalizing) moderates relationship between Avoidant attachment and Detachment.** MASC_correct - correct mentalizing about others.

**S3b Fig. Criterion B. Mentalization about others (undermentalizing) moderates relationship between Avoidant attachment and Detachment.** MASC_minus - undermentalizing about others.

**S3c Fig. Criterion B. Mentalization about others (nonmentalizing) moderates relationship between Avoidant attachment and Detachment.** MASC_brak - nonmentalizing about others

File: S1_Tables (A file encompassing the following tables):

**S1 Table. Regression Analysis Results for the Prediction of Self-Functioning Difficulties.**

**S2 Table. Regression Analysis Results for the Prediction of Interpersonal Functioning Difficulties.**

**S3 Table. Regression Analysis Results for the Prediction of Negative Affect.**

**S4 Table. Regression Analysis Results for the Prediction of Detachment.**

**S5 Table. Regression Analysis Results for the Prediction of Antagonism.**

**S6 Table. Regression Analysis Results for the Prediction of Disinhibition.**

**S7 Table. Regression Analysis Results for the Prediction of Psychoticism.**

